# Family Income and Utilization Disparities for Dental Access of Minority Children: A Cross-Sectional Study

**DOI:** 10.1101/2023.02.07.23285585

**Authors:** David Okuji, Tianqi Wei, Myeonggyun Lee

**Affiliations:** Senior Associate Director, NYU Langone Dental Medicine, Hansjörg Wyss Department of Plastic Surgery, Division of Dental Medicine, NYU School of Medicine, 5800 Third Avenue, 3^rd^ Floor, Brooklyn, NY 11220; Pre-doctoral student, Harvard School of Dental Medicine, 188 Longwood Avenue, Boston, MA 02115; Doctoral student, Division of Biostatistics, Department of Population Health, New York University Grossman School of Medicine, 180 Madison Avenue, Fifth Floor, New York, NY 10016

**Keywords:** child, dentists, health-services-accessibility, systemic-racism, social-class

## Abstract

**Purpose:** Through the use Minorities’ Diminished Return theory, this study aimed to assess the concepts of the theory for access to and utilization of dental care of minority children, compared to White children, when all race/ethnicity groups achieve higher socioeconomic status.

**Methods:** This study was designed cross-sectionally from 21,599 subjects responding to the 2017 National Survey of Children’s Health. The outcome variables of access to and utilization of dental care were compared across Hispanic and non-Hispanic White, Black, Asian, and Multi-race children. Logistic regression models estimated the effects of race/ethnicity on each outcome, with adjustments for child sex, parental education, child age, and income-to-needs ratio.

**Results:** The findings showed that compared to White children, when all racial/ethnic groups increased family income and socioeconomic status, Black and Multi-race children received less health gains in the outcomes of access to and utilization of dental care; and Hispanic children experienced less access to dental care.

**Conclusions:** Minorities’ Diminished Return theory provides evidence of structural barriers which negatively impact the health gains from higher socioeconomic status for access to and utilization of dental care for Black, Hispanic, and Multi-race children. Dentists and policymakers must address systemic racism and structural barriers for oral health equity among all children.

## Introduction

In 2020, the American Medical Association (**AMA**) issued a press release stating, the “AMA Board of Trustees today pledged action to confront systemic racism” and “recognizes that racism in its systemic, structural, institutional, and interpersonal forms is an urgent threat to public health, the advancement of health equity, and a barrier to excellence in the delivery of medical care.” ^1^ “Systemic racism” encompasses whole systems which include social determinants of health such as political, legal, economic, healthcare, educational, and criminal justice system and includes the structures that uphold the systems.^2^ “Structural racism” focuses on the role of structures such as, laws, policies, institutional practices, and entrenched standards that support the system.^3^

From a systemic perspective, despite the growing number of healthcare providers, improvement in medical and oral healthcare, and breakthroughs in medications; racial/ethnic minority groups of children persistently experience multiple disparities across different health fields and have substandard health conditions compared to privileged groups of children.^4,5^ Many serious chronic conditions such as asthma, mental disorder, epilepsy, and heart diseases are more prevalent in children whose household income levels are below the poverty line.^6^ Racial disparities in the outcomes of childhood asthma, such as mortality and hospitalization, are significantly higher in some minority groups of children than White children. Data sets from the National Center for Health Statistics between 2001 and 2010 revealed the asthma occurrence rate in Black children was two times higher than the rate in White children.^7^ Racial disparities for children in the United States (**U**.**S**.) also exist for chronic kidney disease, with end stage kidney diseases more prevalent in Black children compared to White children. The exact reason for those disparities remains unknown, but possible explanations include socioeconomic status (**SES**) and access to care.^8^

Oral health disparities have not been studied as extensively as medical health disparities, but poor oral health has significant influence on the overall body health of children and adults.^9^ The prevalence of untreated dental caries in African American and Caucasian children between ages 3-5 years old was 19% and 11%, respectively. The difference between the prevalence of untreated dental caries in African American and Caucasian teenagers of age 13 to 15 years old was even higher.^9^ Multiple studies have shown that oral health disparities for the African American community are prevalent and influenced by a multitude of factors, including sociocultural context, structure factors, and family experience.^9,10^ Lower income level and cultural perspective can further alter the behavior and perception of seeking treatments. It is essential that oral healthcare professionals understand how certain racial/ethnic groups have a higher incidence of oral diseases and a lower treatment success rate due to those oral health disparities.^11^

Minorities’ Diminished Return (**MDR**) theory is a social epidemiology construct which demonstrates minority populations receive lower increases in health gains compared to White populations when both minority and White families increase family income and socioeconomic status.^12-15^ MDR theory is based upon differential group vulnerability, which hypothesizes that equal resources result in unequal outcomes, with marginalized groups being systemically disadvantaged relative to the dominant group. MDR theory postulates that lower gains for health outcomes of marginalized groups are because their resources generate less tangible health gains than the privileged group.^16^ Although previous studies revealed the effect of MDR theory on unmet dental care need^12^ (**DCN**) and asthma^15^ between White and Black children, there is no evidence regarding the association between MDR theory and the outcomes of access to and utilization of dental care, across other racial minority groups of children.

Therefore, the purpose of this study was to analyze the 2017 National Survey of Children’s Health (**NSCH**) data set^17^ and apply MDR theory to examine the effect of higher income-to-needs ratio, a proxy for SES, for minority families compared to White families for their children’s access to dental care and utilization of dental care. The null hypothesis for the present study was White children and minority children-groups would receive equal health gains for the variables of access to dental care and utilization of dental care when all racial/ethnic groups have protective and higher income-to-needs ratio and SES.

## Method

### Study Design and Setting

The present study was designed as an analytical, observational, cross-sectional model, which examined secondary data from the 2017 NSCH. The collected data related to the physical and mental health of American children from birth to 17 years old in the 50 states plus the District of Columbia. The study analyzed weighted, nationally representative data utilizing the U.S. Census Bureau racial/ethnic group classifications of Hispanic and Non-Hispanic White, Black, Asian, and Multi-race.^17^

### Study Size, Participants, and Data Sources

A total of 59,135 sample households in the 50 states plus the District of Columbia were initially screened to find eligible children based on age. From all the inclusion-eligible children, the caregivers of 21,599 children completed the topical questionnaire about their children’s physical and mental health with language availability in English and Spanish.^17^ Details of the NSCH subject recruitment process and inclusion criteria were accessed from the NSCH Codebooks.^17^

### Variables and Data Measurement

The current study’s outcomes of interest were defined as dichotomous variables indicating an answer of “yes” or “no” to the questions: 1) enabled-access to dental care (**ADC**) measured with a “yes” response to the question, “Did this child have consistent health insurance coverage during the past 12 months?”, 2) utilization of preventive dental care (**UPDC**) measured with a “yes” response to the question, “During the past 12 months, did this child see a dentist or other oral health care provider for preventive dental care, such as check-ups, dental cleanings, dental sealants, or fluoride treatments?”, and 3) utilization of any dental care (**UADC**) measured with a “yes” response to the question, “during the past 12 months, did this child see a dentist or other oral health care provider for any kind of dental or oral health care?”. Note that ADC qualifies “access” to dental care as “enabled-access” to dental care because consistent health insurance coverage only “enables” access to care and does not serve as a direct proxy for access to care.

Additional variables extracted from the NSCH data set included the following parameters: 1) child age as a continuous variable, 2) child sex as a binary variable of Male or Female, 3) child race and ethnicity defined by non-migrant Hispanic (**Hispanic**), non-Hispanic White (**White**), non-Hispanic Black (**Black**), non-Hispanic Asian (**Asian**), and non-Hispanic Multi-race/ethnicity (**Multi-race**) families, 4) parental education defined by “Less than high school,” “High school degree or General Education Diploma (**GED**),” “Some college or technical school,” and “College degree or higher” as an ordinal variable, and 5) income-to-needs ratio, a proxy for SES, as a continuous variable of the “family poverty ratio” from the NSCH data. The family poverty ratio was calculated as a percentage of the ratio of total family income and the family poverty threshold, based upon family size. Note that income-to-”needs” ratio does not imply “dental needs” and instead reflects adequacy of family income compared to family size.

### Statistical Methods and Quantitative Variables

For descriptive statistics, continuous variables were reported with mean and standard deviations and categorical variables were summarized with frequencies and percentages. The descriptive statistics were reported overall in the pooled data. The distributions of variables were compared across different race/ethnicity groups with the analysis of variance (**ANOVA**) test for continuous variables and the Chi-square test for categorical variables.

The logistic regression models were fitted to estimate the effects of race/ethnicity on each outcome, with adjustments for child sex, parental education, child age, and income-to-needs ratio. For each outcome, this study considered two logistic models: Model 1) only outcomes and Model 2) outcomes with the interaction between race/ethnicity and income-to-needs ratio. To investigate the effects of each variable on outcomes within the race/ethnicity group, the analysis additionally conducted the logistic regression analysis, stratified by race/ethnicity.

All statistical tests were two-sided, with Wald type confidence intervals provided, and p<0.05 was considered statistically significant. All analyses were performed in R version 4.0.3 (The R Foundation for Statistical Computing, Vienna, Austria).^18^ Note that the NSCH data set utilized sampling weights for data analysis in the study to attain population-based estimates. To account for the NSCH weights, *‘survey’* R package was used for the analyses.

### Ethics

The NYU Grossman School of Medicine Institutional Review Board reviewed and determined this study (Protocol number 20-01537) did not qualify as human-subjects research.

## Results

Table 1 shows the descriptive analyses and statistically significant inferential associations. Descriptively, the sample population was mostly male (51.2 percent), with mean age of 8.6 years old, 49.6 percent of parents with their highest educational level of a college degree or higher, and mean income-to-needs ratio of 246.17 percent. Inferentially, associations were demonstrated with income-to-needs ratio and parental high school education. Black and Hispanic children had lower socioeconomic levels, reflected by lower income-to-needs ratio, and higher levels of parental high school or General Education Diploma as their highest education level. Among the outcomes, only access to dental care was statistically significant, with Black (9.5 percent) and Hispanic (12.9 percent) children having higher prevalence for gap in insurance coverage. A post-hoc power analysis demonstrated 100% power (1-beta) with alpha set at 0.05.

**Table 1.**
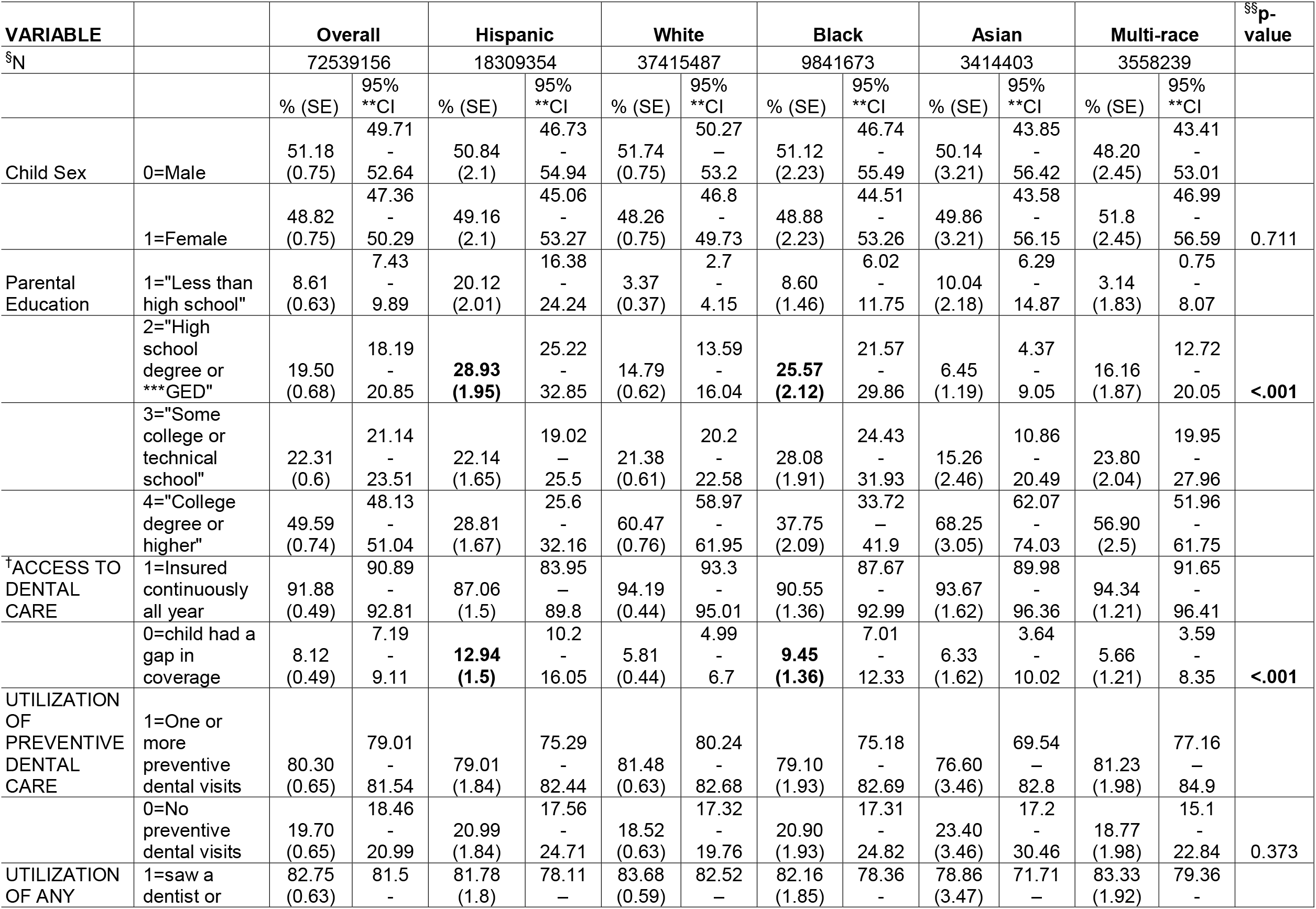
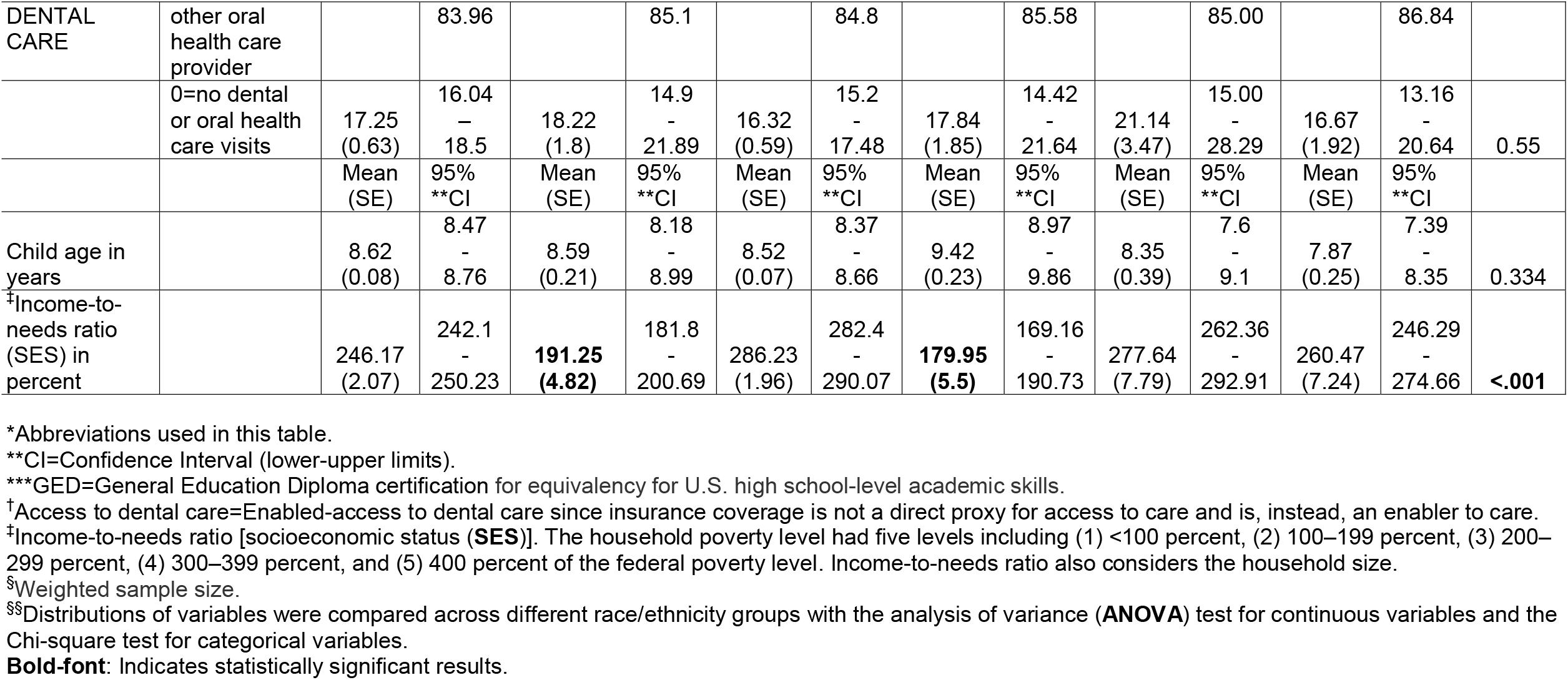
Patient demographics and outcome variables overall, by race*.

Table 2 presents the Model 1 summary of logistic regressions for access to dental care and utilization of dental care, which were estimated in the pooled sample. Model 1 only included the main outcomes. Table 3 presents the Model 2 summary of logistic regressions for access to dental care and utilization of dental care, which were estimated in the pooled sample. Model 2 added the race/ethnicity by income-to-needs ratio interaction. Both Table 2 and Table 3 utilized White race/ethnicity as the reference.

**Table 2.** Pooled Sample Logistic Regressions for Access to dental care, Utilization of preventive dental care, and Utilization of any dental care; with Model 1 (main outcomes)*.

| <sup>†</sup>Access to dental care | Odds Ratio | 95% **CI lower | 95% **CI upper | <sup>§</sup>p |
| --- | --- | --- | --- | --- |
|  | **Model 1 (main outcomes)** |  |  |  |
| Race |  |  |  |  |
| White | 1.000 |  |  |  |
| Hispanic | **0.614** | **0.436** | **0.863** | **0.005** |
| Black | 0.816 | 0.558 | 1.194 | 0.295 |
| Asian | 0.912 | 0.527 | 1.578 | 0.742 |
| Multi-race | 1.086 | 0.675 | 1.747 | 0.733 |
| Child Sex |  |  |  |  |
| Male | 1.000 |  |  |  |
| Female | 1.110 | 0.854 | 1.442 | 0.435 |
| Parental Education |  |  |  |  |
| 1=Less than high school | 1.000 |  |  |  |
| 2=High school degree or GED*** | 1.286 | 0.772 | 2.143 | 0.334 |
| 3=Some college or technical school | 1.430 | 0.881 | 2.321 | 0.148 |
| 4=College degree or higher | **2.714** | **1.538** | **4.791** | **0.001** |
| Child Age | 0.979 | 0.954 | 1.005 | 0.118 |
| Income-to-needs ratio (SES)<sup>‡</sup> | 1.001 | 1.000 | 1.003 | 0.052 |
| **Utilization of preventive dental care** | **Odds Ratio** | **95% \*\*CI lower** | **95% \*\*CI upper** | **<sup>§</sup>p** |
|  | **Model 1 (main outcomes)** |  |  |  |
| Race |  |  |  |  |
| White | 1.000 |  |  |  |
| Hispanic | 1.136 | 0.895 | 1.442 | 0.294 |
| Black | 0.890 | 0.668 | 1.187 | 0.428 |
| Asian | 0.707 | 0.471 | 1.061 | 0.094 |
| Multi-race | 1.137 | 0.861 | 1.502 | 0.364 |
| Child Sex |  |  |  |  |
| Male | 1.000 |  |  |  |
| Female | 0.997 | 0.843 | 1.180 | 0.973 |
| Parental Education |  |  |  |  |
| 1=Less than high school | 1.000 |  |  |  |
| 2=High school degree or GED*** | **1.625** | **1.060** | **2.491** | **0.026** |
| 3=Some college or technical school | **1.852** | **1.231** | **2.786** | **0.003** |
| 4=College degree or higher | **3.091** | **2.089** | **4.574** | **0.000** |
| Child Age | **1.198** | **1.170** | **1.226** | **0.000** |
| Income-to-needs ratio (SES)<sup>‡</sup> | 1.000 | 1.000 | 1.001 | 0.288 |
| **Utilization of any dental care** | **Odds Ratio** | **95% \*\*CI lower** | **95% \*\*CI upper** | **<sup>§</sup>p** |
|  | **Model 1 (main outcomes)** |  |  |  |
| Race |  |  |  |  |
| White | 1.000 |  |  |  |
| Hispanic | 1.140 | 0.884 | 1.471 | 0.313 |
| Black | 0.884 | 0.648 | 1.206 | 0.438 |
| Asian | 0.689 | 0.448 | 1.062 | 0.091 |
| Multi-race | 1.135 | 0.850 | 1.515 | 0.390 |
| Child Sex |  |  |  |  |
| Male | 1.000 |  |  |  |
| Female | 0.958 | 0.799 | 1.148 | 0.642 |
| Parental Education |  |  |  |  |
| 1=Less than high school | 1.000 |  |  |  |
| 2=High school degree or GED*** | **1.689** | **1.057** | **2.698** | **0.028** |
| 3=Some college or technical school | **2.032** | **1.300** | **3.176** | **0.002** |
| 4=College degree or higher | **3.213** | **2.102** | **4.912** | **0.000** |
| Child Age | **1.230** | **1.197** | **1.265** | **0.000** |
| Income-to-needs ratio (SES)<sup>‡</sup> | 1.000 | 0.999 | 1.001 | 0.763 |

Notes:
\*Abbreviations used in this table.
\*\*CI=Confidence Interval.
\*\*\*GED=General Education Diploma certification for equivalency for U.S. high school-level academic skills.
<sup>†</sup>Access to dental care=Enabled-access to dental care since insurance coverage is not a direct proxy for access to care and is, instead, an enabler to care.
<sup>‡</sup>Income-to-needs ratio [socioeconomic status (**SES**)]. The household poverty level had five levels including (1) <100 percent, (2) 100–199 percent, (3) 200–299 percent, (4) 300–399 percent, and (5) 400 percent of the federal poverty level. Income-to-needs ratio also considers the household size.
<sup>§</sup>Logistic regression models were fitted to estimate the effects of race/ethnicity on each outcome, with adjustments for child sex, parental education, child age, and income-to-needs ratio. For each outcome, Model 1 only considered main outcomes. To investigate the effects of each variable on outcomes within the race/ethnicity group, the analysis additionally conducted the logistic regression analysis, stratified by race/ethnicity.
**Bold-font:** Indicates statistically significant results.

**Table 3.** Pooled Sample Logistic Regressions for Access to dental care, Utilization of preventive dental care, and Utilization of any dental care; with Model 2 (interaction between outcome of race and income-to-needs ratio)*.

| <sup>†</sup> Access to dental care | Odds Ratio | 95% **CI lower | 95% **CI upper | <sup>§</sup>p |
| --- | --- | --- | --- | --- |
| **Model 2 (race by income-to-needs ratio interaction)** |  |  |  |  |
| Race |  |  |  |  |
| White | 1.000 |  |  |  |
| Hispanic | 1.303 | 0.728 | 2.333 | 0.373 |
| Black | 1.573 | 0.887 | 2.791 | 0.121 |
| Asian | 0.821 | 0.322 | 2.091 | 0.679 |
| Multi-race | 0.987 | 0.401 | 2.428 | 0.977 |
| Child Sex |  |  |  |  |
| Male | 1.000 |  |  |  |
| Female | 1.105 | 0.852 | 1.432 | 0.452 |
| Parental Education |  |  |  |  |
| 1=Less than high school | 1.000 |  |  |  |
| 2=High school degree or GED*** | 1.356 | 0.821 | 2.239 | 0.234 |
| 3=Some college or technical school | 1.537 | 0.947 | 2.493 | 0.082 |
| 4=College degree or higher | **2.781** | **1.597** | **4.843** | **0.000** |
| Child Age | 0.981 | 0.955 | 1.006 | 0.140 |
| Income-to-needs ratio (SES)<sup>‡</sup> | **1.003** | **1.002** | **1.004** | **0.000** |
| Income-to-needs ratio X Race |  |  |  |  |
| status:Hispanic | **0.996** | **0.994** | **0.999** | **0.001** |
| status:Black | **0.997** | **0.994** | **0.999** | **0.015** |
| status:Asian | 1.001 | 0.998 | 1.004 | 0.636 |
| status:Multi-race | 1.001 | 0.997 | 1.005 | 0.654 |
| **Utilization of preventive dental care** | **Odds Ratio** | **95% \*\*CI lower** | **95% \*\*CI upper** | **§p** |
|  | **Model 2 (race by income-to-needs ratio interaction)** |  |  |  |
| Race |  |  |  |  |
| White | 1.000 |  |  |  |
| Hispanic | 1.425 | 0.904 | 2.248 | 0.127 |
| Black | **1.755** | **1.082** | **2.848** | **0.023** |
| Asian | 1.389 | 0.556 | 3.470 | 0.481 |
| Multi-race | **2.063** | **1.138** | **3.742** | **0.017** |
| Child Sex |  |  |  |  |
| Male | 1.000 |  |  |  |
| Female | 0.998 | 0.844 | 1.179 | 0.979 |
| Parental Education |  |  |  |  |
| 1=Less than high school | 1.000 |  |  |  |
| 2=High school degree or GED*** | **1.627** | **1.066** | **2.483** | **0.024** |
| 3=Some college or technical school | **1.874** | **1.247** | **2.818** | **0.003** |
| 4=College degree or higher | **3.101** | **2.100** | **4.580** | **0.000** |
| Child Age | **1.198** | **1.171** | **1.227** | **0.000** |
| Income-to-needs ratio (SES)<sup>†</sup> | **1.001** | **1.001** | **1.002** | **0.000** |
| Income-to-needs ratio X Race |  |  |  |  |
| status:Hispanic | 0.999 | 0.997 | 1.001 | 0.371 |
| status:Black | **0.997** | **0.995** | **0.999** | **0.001** |
| status:Asian | 0.998 | 0.994 | 1.001 | 0.114 |
| status:Multi-race | **0.998** | **0.996** | **1.000** | **0.024** |
| **Utilization of any dental care** | **Odds Ratio** | **95% \*\*CI lower** | **95% \*\*CI upper** | **§p** |
|  | **Model 2 (race by income-to-needs ratio interaction)** |  |  |  |
| Race |  |  |  |  |
| White | 1.000 |  |  |  |
| Hispanic | 1.300 | 0.800 | 2.113 | 0.289 |
| Black | **1.808** | **1.071** | **3.054** | **0.027** |
| Asian | 1.147 | 0.432 | 3.049 | 0.783 |
| Multi-race | **2.091** | **1.133** | **3.858** | **0.018** |
| Child Sex |  |  |  |  |
| Male | 1.000 |  |  |  |
| Female | 0.959 | 0.801 | 1.148 | 0.647 |
| Parental Education |  |  |  |  |
| 1=Less than high school | 1.000 |  |  |  |
| 2=High school degree or GED*** | **1.663** | **1.048** | **2.641** | **0.031** |
| 3=Some college or technical school | **2.026** | **1.297** | **3.165** | **0.002** |
| 4=College degree or higher | **3.180** | **2.086** | **4.850** | **0.000** |
| Child Age | **1.231** | **1.197** | **1.265** | **0.000** |
| Income-to-needs ratio (SES)<sup>†</sup> | **1.001** | **1.000** | **1.002** | **0.011** |
| Income-to-needs ratio X Race |  |  |  |  |
| status:Hispanic | 1.000 | 0.998 | 1.002 | 0.753 |
| status:Black | **0.996** | **0.994** | **0.999** | **0.001** |
| status:Asian | 0.998 | 0.995 | 1.001 | 0.270 |
| status:Multi-race | **0.998** | **0.996** | **1.000** | **0.026** |

Notes:
\*Abbreviations used in this table.
\*\*CI=Confidence Interval.
\*\*\*GED=General Education Diploma certification for equivalency for U.S. high school-level academic skills.
†Access to dental care=Enabled-access to dental care since insurance coverage is not a direct proxy for access to care and is, instead, an enabler to care.
‡Income-to-needs ratio [socioeconomic status (**SES**)]. The household poverty level had five levels including (1) <100 percent, (2) 100–199 percent, (3) 200–299 percent, (4) 300–399 percent, and (5) 400 percent of the federal poverty level. Income-to-needs ratio also considers the household size.
§Logistic regression models were fitted to estimate the effects of race/ethnicity on each outcome, with adjustments for child sex, parental education, child age, and income-to-needs ratio. For each outcome, Model 2 considered outcomes with the interaction between race/ethnicity and income-to-needs ratio. To investigate the effects of each variable on outcomes within the race/ethnicity group, the analysis additionally conducted the logistic regression analysis, stratified by race/ethnicity.
**Bold-font:** Indicates statistically significant results.

Statistically significant results in Table 2, Model 1 show for ADC, with all other covariates fixed, Hispanic children tended to have lower ADC than White children (Odds Ratio (**OR**) 0.614, 95 percent confidence interval (**95% CI**) 0.436 to 0.863) (*P*=0.005). Also with fixed covariates, parents with a college degree or higher had higher ADC (OR 2.714 [95% CI 1.538 to 4.791] [*P*=.0.001]). For UPDC and UADC, parents with lower education levels have lower probability of UPDC and UADC; the higher the age, the higher the UPDC (OR 1.198 [95% CI 1.170 to 1.226] [*P*=.0.000]) and UADC (OR 1.230 [95% CI 1.197 to 1.265] [*P*=.0.000]).

Statistically significant results in Table 3, Model 2 demonstrate for higher parental college education, the higher ADC, UPDC, and UADC. The most critical analysis for the effect of increased income-to-need ratio (SES) is shown in Table 3, Model 2 highlighted in gray color. An interaction between race/ethnicity and income-to-needs ratio was identified, which estimated a statistically significant and low magnitude trend for the protective effect of the income-to-needs ratio for White children than 1) Black children for access to dental care (OR 0.997 [95% CI 0.994 to 0.999] [*P*=.0.015]), utilization of preventive dental care (OR 0.997 [95% CI 0.995 to 0.999] [*P*=.0.001]), and utilization of any dental care (OR 0.996 [95% CI 0.994 to 0.999] [*P*=.0.001]), 2) Hispanic children for access to dental care (OR 0.996 [95% CI 0.994 to 0.999] [*P*=.0.001]), and 3) Multi-race children for utilization of preventive dental care (OR 0.998 [95% CI 0.996 to 1.000] [*P*=.0.024]) and utilization of any dental care (OR 0.998 [95% CI 0.996 to 1.000] [*P*=.0.026]).

In sum, although all racial/ethnic groups benefitted from the protective effect of higher income and SES; Black, Hispanic, and Multi-race children had significantly lower protection, of low magnitude, than White children. Specifically, Black and Hispanic children had lower ADC; and Black and Multi-race children had lower UPDC and UADC. There were no significant differences between Asian and White children for access to dental care and utilization of dental care.

## Discussion

### Findings and Hypotheses

The results of the current study, using 2017 NSCH data, rejected the null hypothesis, and demonstrated that MDR theory is applicable in the health outcomes of access to dental care and utilization of dental care for minority children, when comparing higher SES parity between White and minority populations.

This present study’s findings showed a trend of low magnitude significance for disparities between White and minority children for access to and utilization of dental care, even when all children gain higher SES. Black children received less health gains from their higher SES compared to White children in the outcomes of access to dental care and utilization of dental care. Hispanic children experienced less access to dental care compared to White children despite having higher SES. Multi-race children had unequal gains from their SES compared to White children for the outcomes of utilization of dental care. Asian children did not exhibit less health gains from higher protective income-to-needs ratio compared to White children.

### Comparison to Literature

The finding that Black children receive less protective effect of SES for access to preventive dental care is externally validated in a previously study by Assari, which showed that Black children endure more unmet “dental care need” than White children despite the two racial/ethnic groups having similar family income-to-needs ratio.^12^ The present study’s authors believe Assari inaccurately defined unmet “dental care need” as a “no” response, using a single item variable by asking, “During the past 12 months, did this child see a dentist or other oral health care provider for preventive dental care, such as check-ups, dental cleanings, dental sealants, or fluoride treatments?” The more accurate definition for this question, with a “yes” response, should be “utilization of preventive dental care” as specified within this present study.

This present study expanded MDR to the outcomes of access to and utilization of dental care, and demonstrated Black children have less access to dental care and utilization of dental care when they have similar SES as White children. This could potentially contribute to the socio-cultural influence on the perception of oral diseases. A study regarding the connection of culture and behavior demonstrated that African Americans do not actively seek oral health care because they think dental caries are not a health issue.^11^ There are also other studies that show MDR exist in various health outcomes such as smoking behavior, chronic health condition, mortality, mental and physical health when the researchers compared the income and education level of Black population and White population.^19-25^ In addition, a 2021 study concluded racial discrimination perceived by Black and Hispanic adults, with the same risk factors, in part explains racial disparities for dental utilization.^26^

Previous studies demonstrate the effects of MDR in multiple medical health domains. This study’s additional findings in oral health access and utilization, further emphasize that health inequality between Black children and White children is not necessarily due to individual and culture differences, with societal and structure systems as the contributing barriers which Black families face when they try to achieve the similar living standard as the White families.^13-15,25,27^

This study is the first to evaluate the association of oral health care and MDR in Hispanic children. Past literature suggests the cause of poor oral health in Hispanic children, especially in preventive dental care services, was family income and neighborhood environment. Thus, the policy-making direction was primarily focused on expanding pediatric dental coverage and oral hygiene education. While those causes might be true, this study’s finding revealed that Hispanic children would not achieve adequate dental care services even if family incomes increase to the same level as the White families. This, again, indicates that structural racism might be a societal barrier preventing Hispanic children from gaining equal oral health care.^28-30^

There are few studies about racial disparities in oral health for multi-race children, possibly due to insufficient sample size in national databases.^5,31^ Other studies showed that multi-race children had significantly higher incidence of missing preventive dental visits in the past 12 months and demonstrated that the low frequency of dental visits is not primarily due to family income but instead to structural racism. ^4,5^

### Strengths and Limitations

The main strength of this study is it is the first to apply MDR theory between multiple racial/ethnic groups for the outcomes of access to and utilization of dental care. This study’s wide range of racial population strengthens the definition of MDR by extending to all minority racial/ethnic groups, rather than solely restricted to the Black population. The use of access to and utilization of dental care as dependent variables also enhance the measurements of how children of minority populations receive their oral health care. In addition, the results of this study are generalizable with its large and diverse sample, high level of statistical power, and U.S. government data source which was nationally representative for children’s health.

However, the interpretation of this study must be carefully considered with appropriate limitations. Confirmation bias may be a limitation given the low magnitude of the significant differences in odds ratios shown in Table 2 and Table 3. This research study does not determine the cause-and-effect relationship between any variables used throughout the process of research. As well, the majority responders of the NSCH program were parents, and their interpretations of their children’s health might not be fully accurate. Since the entire data were extracted from a questionnaire-based survey, the question structures and word choices might affect the judgment of the responders which might also depend upon their family cultures and social experiences. Finally, the data were collected in 2017 and thus should be used with caution when applying the findings to current economic and health outcome conditions.

### Significance and Implications

It is crucial that policymakers realize that equalizing SES might not completely resolve oral health disparities among minority children. The oral health disparities which exist in society are rooted deeply inside the structure of society, health care system, discrimination between race/ethnicity and life stress.^32-34^

An example of how the structure of healthcare financing creates oral health disparities is the Medicaid system. According to 2016 data, nearly twice as many physicians (69%) participated as Medicaid providers compared to dentists (38%), despite nearly equal reimbursement rates.^35^ Hence, there are fewer dentist providers compared to physicians.

Medicaid enrollment data show 61.1% of beneficiaries are Hispanic, Black, Asian, or a non-White race or ethnicity.^36^ Juxtaposed data indicate White-dentists composed 70.2% of the dentist workforce, with Asian-, Hispanic-, and Black-dentists representing 18%, 5.9%, and 3.8%, respectively.^37^ Thus, there is structural misalignment between the racial/ethnic composition of Medicaid enrollees and dentist-providers. Structural barriers, which explain why a large majority of dentists do not participate as Medicaid providers; include low reimbursement rates, administrative inefficiencies impacting Medicaid providers, high number and prevalence of missed appointments by Medicaid beneficiaries, and a shortage of dentists from diverse backgrounds.^34^ Medicaid policymakers should address these types of misalignments by removing structural barriers for improved oral health in children.

In 2003 the Institute of Medicine (now known as the National Academy of Medicine) published its seminal report, “Unequal Treatment: Confronting Racial and Ethnic Disparities in Health Care.”^38^ The report identified health systems-level variables such as the ways in which systems are organized, financed, accessed, and utilized might have varying impacts on patient care racial/ethnic minorities. Examples of these variables include sub-optimal communication between providers and patients with low levels of English-language proficiency; time pressures on healthcare providers, with insufficient time to accurately assess minority patients; and disparities in geographic availability of healthcare institutions between minority and majority groups, due to differences in neighborhood income levels.^37^ Hence, future policies regarding oral health care should address the above-noted systemic factors by removing social barriers, mitigating discrimination, and re-designing a more comprehensive and equitable health care system. Oral health clinicians should be aware that minority groups of children who have worse oral health maintenance may likely be due to systemic and structural barriers.

### Future Studies

To strengthen the evidence for MDR theory, future studies might be designed with prospective and longitudinal constructs which generate predictive statistical analyses. This study’s findings, generated by the 2017 NSCH data, did not include how MDR theory might be applied to specific clinical outcomes. Hence, future studies might also explore more recent NSCH data sets to learn how MDR theory might apply to oral health outcomes, such as dental caries or other oral health problems, and be compared among racial/ethnic groups.

## Conclusion

Based on this study’s results, the following conclusions can be made:

1. Evidenced by logistical regression analysis, the concepts of Minorities’ Diminished Return theory were supported for the outcomes of access to and utilization of dental care among racial/ethnic groups of Black, Hispanic, and Multi-racial minority children.
2. Dentists and policymakers should address systemic racism and structural barriers for oral health equity among all children through increased access to and utilization of dental care for minority children within dental practices by, for example, welcoming and providing care to children with Medicaid benefits.

## Data Availability

All data produced in the present study are available upon reasonable request to the authors

https://www.childhealthdata.org/learn-about-the-nsch/nsch-codebooks

## Acknowledgments

The authors thank the Hansjörg Wyss Department of Plastic Surgery, NYU Langone Health for funding; Veronica Brandley, DMD, Paulina Nguyen, DDS, and Regina Nguyen, DDS who assisted this project during residency training; and Jay Balzer, DMD, MPH, Amy Kim, DDS, and Adolfina Polk, DDS who provided manuscript review.

